# Systematic evaluation of the environmental effect on depressive symptoms in late adolescence and early adulthood: exposome-wide association study and twin modeling

**DOI:** 10.1101/2023.03.27.23287786

**Authors:** Zhiyang Wang, Stephanie Zellers, Alyce M. Whipp, Marja Heinonen-Guzejev, Maria Foraster, Jordi Júlvez, Irene van Kamp, Jaakko Kaprio

**Affiliations:** Institute for Molecular Medicine Finland, Helsinki Institute of Life Science, University of Helsinki, Helsinki, Finland; Department of Public Health, University of Helsinki, Helsinki, Finland; PHAGEX Research Group, Blanquerna School of Health Science, Universitat Ramon Llull (URL), Barcelona, Spain; ISGlobal, Parc de Recerca Biomèdica de Barcelona (PRBB), Barcelona, Spain; Universitat Pompeu Fabra (UPF), Barcelona, Spain; CIBER Epidemiología y Salud Pública (CIBEREsp), Madrid, Spain; Clinical and Epidemiological Neuroscience (NeuroÈpia), Institut d’Investigació Sanitària Pere Virgili (IISPV), Reus, Spain; Centre for Sustainability, Environment and Health, National Institute for Public Health and the Environment (RIVM, Netherlands), Bilthoven, the Netherlands

**Author notes:** Corresponding author: Jaakko Kaprio; address: Institute for Molecular Medicine, University of Helsinki, PL 20 (Tukholmankatu 8), FI-00014, Helsinki, Finland.

## Abstract

The exposome represents the totality of environmental effects, but systematic evaluation between it and depressive symptoms is scant. We sought to comprehensively identify the association of the exposome with depressive symptoms in late adolescence and early adulthood and determine genetic and environmental covariances between them. Based on the FinnTwin12 cohort (3025 participants in young adulthood and 4127 at age 17), the exposome-wide association study (ExWAS) design was used to identify significant exposures from 12 domains. Bivariate Cholesky twin models were fitted to an exposome score and depressive symptoms. In ExWASes, 29 and 46 exposures were significantly associated with depressive symptoms in young adulthood and at age 17, respectively, and familial exposures were the most influential. Twin models indicated considerable genetic and environmental covariances between the exposome score and depressive symptoms with sex differences. The findings underscore the systematic approach of the exposome and the consideration of relevant genetic effects.

## Introduction

Depressive symptoms are a type of chronic mental health condition with complex etiology, and major depressive disorder (MDD) is the clinical disorder diagnosed when depressive symptoms reach a threshold of severity and duration. Depressive symptoms and MDD lead to a serious public health burden. The updated Global Burden of Diseases study showed that the age-standardized prevalence of MDD was 4% (3951 per 100,000 people) in Western Europe, higher than the global level, and underlined the heavy burden on people aged between 15 and 24^1^. Among adolescents, a 2021 systematic review indicated that the pooled prevalence of self-reported depressive symptoms was 34% and of MDD was 5% from the studies between 2001 to 2020, and the prevalence is increasing^2^. The COVID-19 pandemic exacerbated the already growing trend of hardship. Given a growing body of evidence on the environmental effect on depressive symptoms and MDD^3,4^, more systematic investigation is urgently needed, especially among youth.

The concept of the “exposome” was raised in 2005, which depicts the dynamic totality of the environment that an individual experiences^5^. The exposome is divided into three parts: specific external, general external, and internal exposomes, and the external exposome could be further subdivided into the familial, social, built exposome, and so on. Instead of studying a single or small group of exposures, an exposome study aims to investigate the overall effect of the environment, while, unavoidably, complexities like interaction or ubiquity increase the difficulty^6^. An exposome (environmental, exposure) -wide association study (ExWAS), likes other “WAS” studies, denotes an agnostic and systematic method for hypothesis-generating, which is comparatively appropriate to the exposome’s spatiotemporal variabilities and multi-level structure^7^. Several ExWAS studies have targeted mental health^8–10^, and Choi et al.^11^ used clinically significant incident depression as the outcome and identified multiple modifiable factors. As the early warning sign of MDD, focusing on depressive symptoms in adolescence or young adulthood could be easier to guide translational intervention as early as possible, which would be more cost-effective.

Despite the benefits of the exposome approach, there are some other hindrances. First, under the current technique, we cannot measure every possible exposure (far from reaching “1-genome”), and the exposome keeps updating, expanding, and enriching. Moreover, some studies have emphasized exposures’ non-genetic properties, which ignores how the environment interacts with genetics through multiple mechanisms among many traits including depression^12,13^. Medda and colleagues, based on the Italian Twin Registry, demonstrated the substantial genetic role in exogenous metallomics, where the estimations of standardized genetic variance, as a proportion of total variance of the measured exposures, ranged from 0.15 (Arsenic) to 0.79 (Zinc)^14^. As a natural experiment, twin and family studies provide one method to evaluate genetic and environmental relationships between traits and exposures. This design decomposes the variance of traits into additive genetic (A), domaint genetic (D), common environmental (C), and unique environmental (E) components, which contain the distinct features of the exposome as the overall environmental effect. Such indirect evidence of genetic effects based on genetic relationships of family members is an efficient way to demonstrate the presence (or lack of) genetic effects. Thus, the combination of exposome and twin studies could advance our knowledge of the complexities between genes and environments, improve our understanding of existing deficiencies in exposome measures, and produce further research questions. A natural extension is then to include measured genotypes, either targeting specific genes such as those involved in the metabolism of external compounds or more broad-based genome-wide approaches to derive polygenic scores of genetic susceptibility.

In this study, based on the FinnTwin12 cohort, we aim to: 1) comprehensively and systematically determine exposures that are significantly associated with depressive symptoms and MDD in late adolescence and early adulthood through three ExWASes and 2) estimate to what extent the exposome score and depressive symptoms share the same genetic and environmental risk factors.

## Results

### Characteristics of the study, participants, and exposures

Figure 1 shows the flowchart of the analysis pipeline, which consisted of three ExWASes and the following bivariate twin modeling. Based on the FinnTwin12 cohort, there were 3025, 1236, and 4127 individual twins included in three separate ExWASes with the outcomes of general behavior inventory (GBI) score in young adulthood (primary), the incidence of MDD in young adulthood, and GBI score at age 17, respectively. The characteristics of each ExWAS are shown in Table 1.

**Figure 1:**
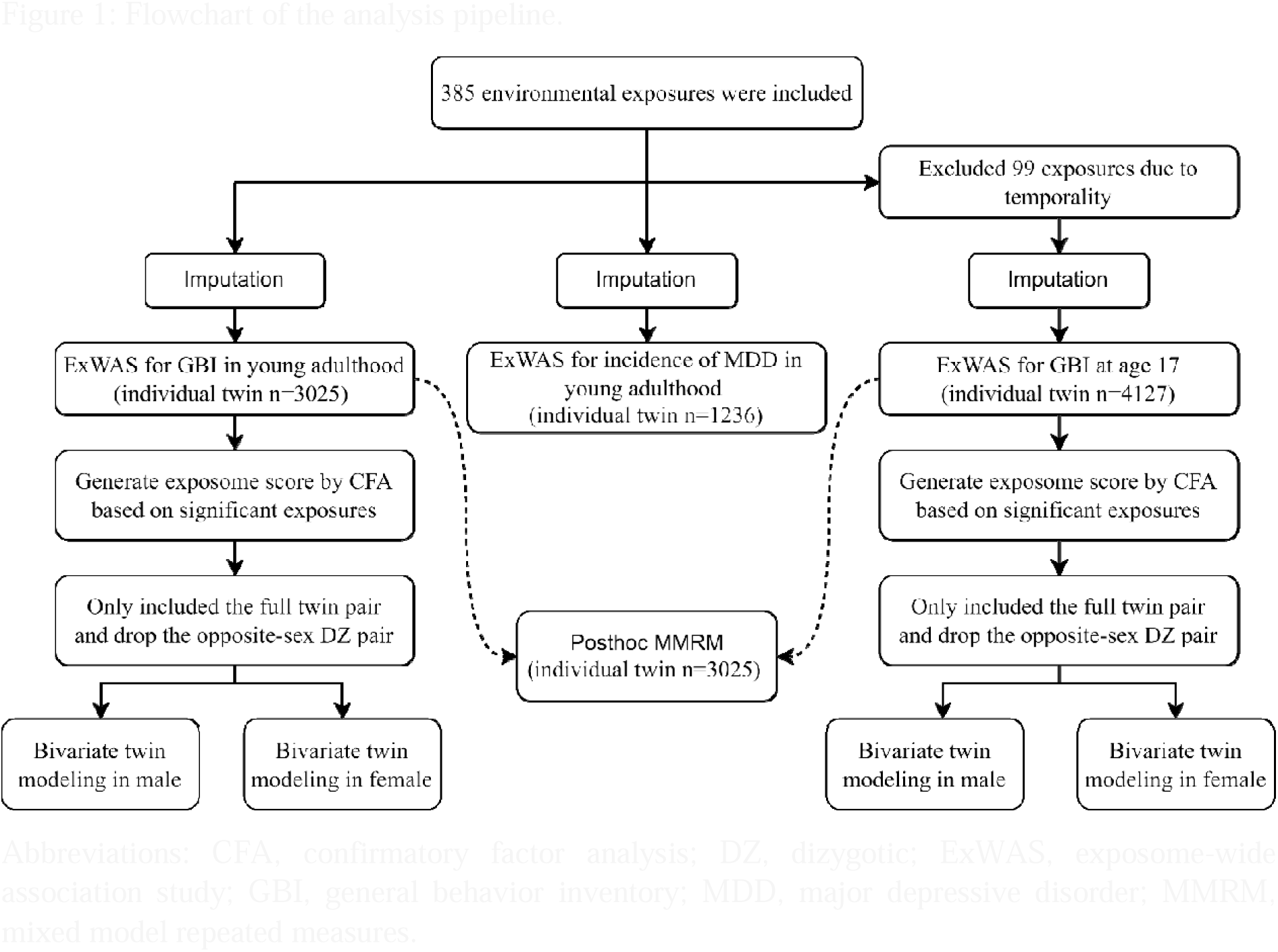
Flowchart of the analysis pipeline. Abbreviations: CFA, confirmatory factor analysis; DZ, dizygotic; ExWAS, exposome-wide association study; GBI, general behavior inventory; MDD, major depressive disorder; MMRM, mixed model repeated measures.

**Table 1:** Characteristics of ExWASes.

| **Outcome** | **Number of individual twins** | **Number of exposures** | **Number of P values** | **Significant threshold ($-\log_{10}(\text{P value})$)** |
| --- | --- | --- | --- | --- |
| GBI in young adulthood | 3025 | 385 | 501 | 3.51 |
| Incidence of MDD in young adulthood | 1236 | 385 | 501 | 3.47 |
| GBI at age 17 | 4127 | 286 | 394 | 3.44 |

For individual twins included in ExWASes of all outcomes (Table 2), the majority were female and from dizygotic (DZ) pairs, and their parental education levels were limited (less than high school). At age 17, 25.3% of individual twins reported being current smokers and 82.6% were full-time students and not working. In young adulthood, 25.4% of individual twins reported that they were currently smoking and 51.4% had a full-time job. The mean GBI scores at age 17 and in young adulthood were 5.0 (SD: 4.9) and 4.4 (SD: 4.7), respectively, and the two measures correlated with 0.49. The incidence of lifetime MDD in young adulthood was 12.3%.

**Table 2:**
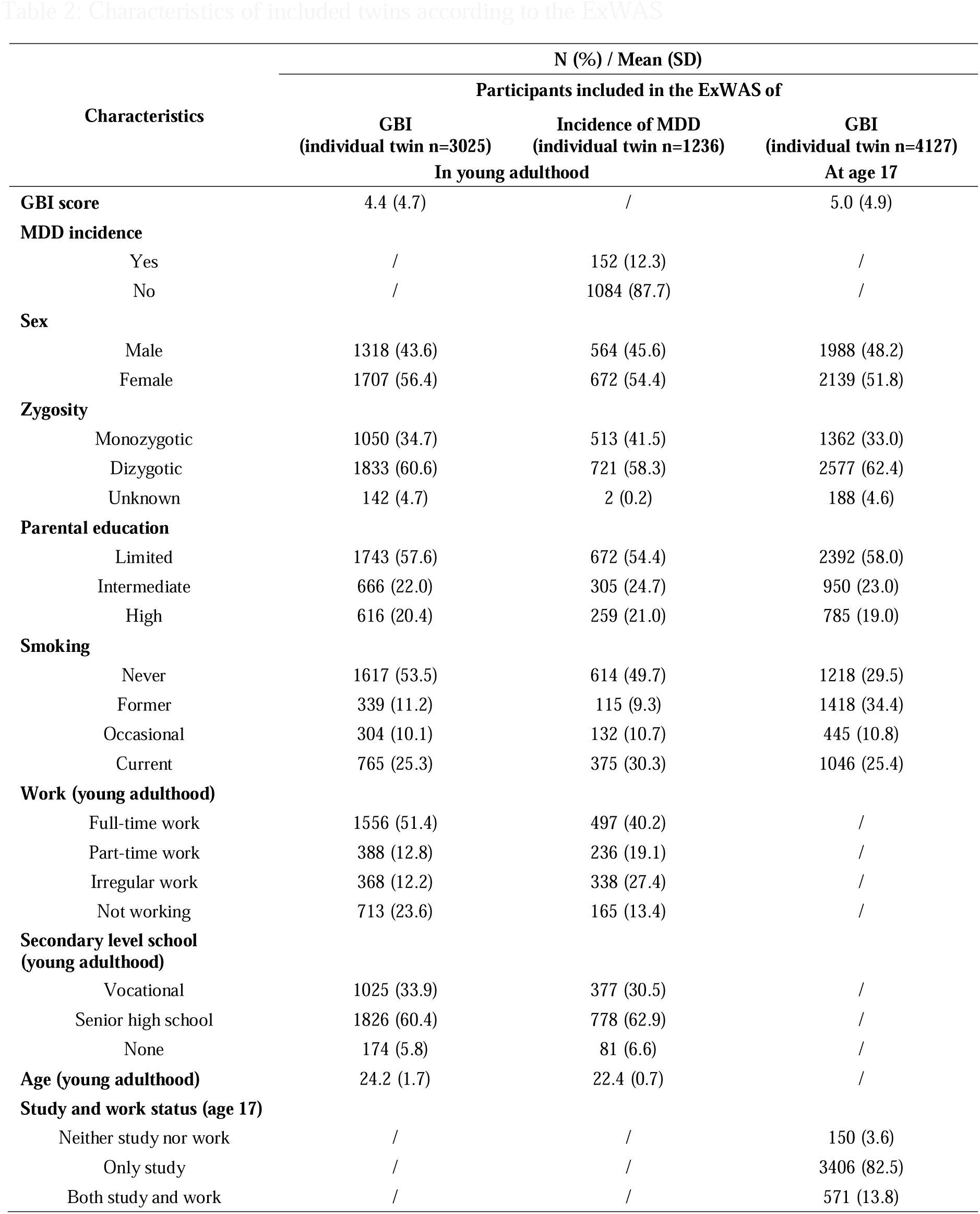
Characteristics of included twins according to the ExWAS.

Exposures’ codenames, description, and statistics based on twins included in the ExWAS of GBI in young adulthood (before imputation) are presented in Supplementary Table 1. There are 12 domains of exposures, colored in the following plots: air pollution, building, blue and green spaces, population density, geocoordinates, prenatal exposures, passive smoking, family and parents, friend and romantic relationships, school and teachers, stressful life events, and social indicators. In principal component analysis (PCA), the first principal component (PC1) only attributed 10.93% and 10.66% to the total variability of all included exposures in young adulthood and age 17, respectively (Supplementary Figure 1). From the scatter plots of PC1 and PC2, we identified some potential clusters of exposures from domains of building, blue and green spaces, and social indicators via visual assessment.

### ExWAS of log-transformed GBI score and incidence of MDD in young adulthood

The adjusted coefficient and -*log*_10_(*P value*) of all exposures included for both adult outcomes are presented in Supplementary Table 2. There were 40 significant P values in 29 exposures, which were associated with log-transformed GBI score in young adulthood, identified from 385 exposures (Figure 2A). There were 24, 2, and 3 exposures belonging to the domains of family and parents, friend and romantic relationships, and school and teachers, respectively. For the most protective exposure, compared to twins who felt their home environment was completely unfair, quite unfair, or somewhat unfair at age 17 (unfair_A17), twins who felt it was not at all unfair at age 17 were associated with a 0.40 lower log-transformed GBI score (95% CI: −0.50, −0.31) (Figure 2B). For the most harmful exposure, compared to twins who were completely satisfied with their relationship with friends at age 14 (sat_friend_A14), twins who felt somewhat satisfied, mainly not satisfied, or not at all satisfied at age 14 were associated with a 0.42 higher log-transformed GBI score (95% CI: 0.29, 0.55) (Figure 2B). In contrast, none of the exposures showed a significant association with MDD (Supplementary Figure 2).

**Figure 2:**
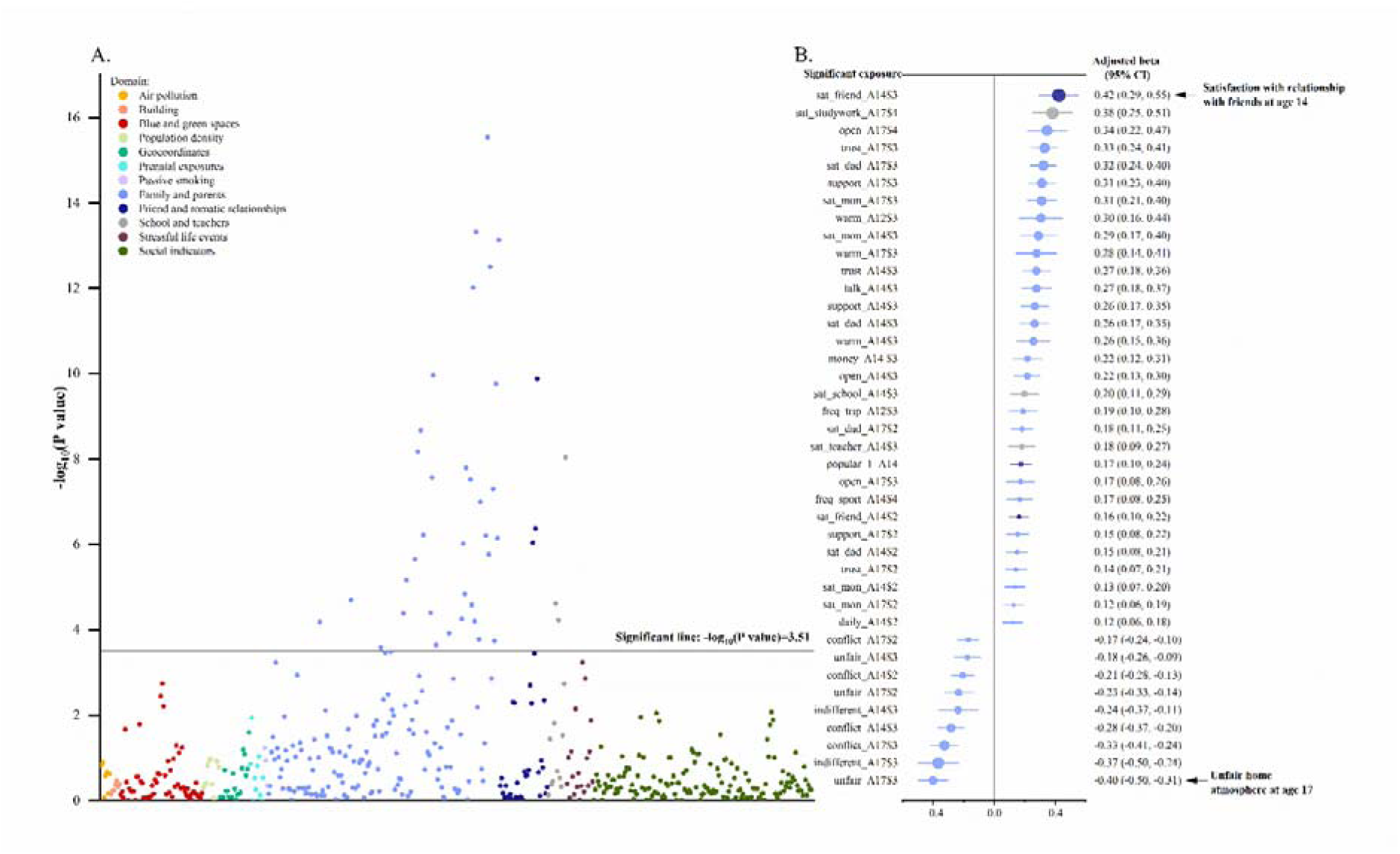
Association results between exposure and log-transformed GBI score in young adulthood, adjusted for covariates ^a^ ^a^ Panel A is a Manhattan association plot for exposures in relation to log-transformed GBI score in young adulthood. The y-axis is showing statistical significance as –*log*_10_(*P value*). Panel B presents the adjusted beta for significant exposures in descending order (from harmful to protective). The color legend applies to both Panel A (Manhattan association plot) and B (forest plot). The adjusted covariates were: sex, zygosity, parental education, smoking in young adulthood, work status in young adulthood, secondary level school in young adulthood, and age when twins provided the GBI assessment in young adulthood.

### ExWAS of log-transformed GBI score at age 17

The adjusted coefficient and -log_10_(P value) for the age 17 outcome were presented in Supplementary Table 2. There were 71 significant P values in 46 exposures, which were significantly associated with log-transformed GBI score, identified from 286 exposures (Supplementary Figure 3A). There were 32, 6, 4, and 4 exposures belonging to the domains of family and parent, friend and romantic relationship, school and teachers, and stressful life events, respectively. For the most harmful exposures, compared to twins who were completely satisfied with their success at work or studies at age 17 (sat_studywork_A17), twins who felt mainly not satisfied, or not at all satisfied at age 17 were associated with a 0.65 higher log-transformed GBI score (95% CI: 0.55, 0.74) (Supplementary Figure 3B). For the most protective exposure, the same as the result in young adulthood, compared to twins who felt their home environment was completely unfair, quite unfair, or somewhat unfair at age 17 (unfair_A17), twins who felt it was not at all unfair at age 17 were associated with a 0.50 lower log-transformed GBI score (95% CI: - 0.57, −0.43) (Supplementary Figure 3B). There are 27 exposures that are significantly associated with both log-transformed GBI scores in young adulthood and at age 17, and of 22 exposures belong to the domain of family and parents.

### Twin modeling of depressive symptoms with exposome scores

Before the bivariate modeling, the best-fit univariate AE model (had the lowest Akaike information criterion (AIC) compared to ADE and E models) indicated E explained 61% of the variance of depressive symptoms in males and 45% in females at age 17, and the numbers slightly reduced to 59% and 42% in young adulthood, respectively (Supplementary Table 3). The exposome score was created by confirmatory factor analysis (CFA) based on the significant exposures from ExWASes. The standardized root mean square residual (SRMR) of models in young adulthood and age 17 were 0.100 and 0.078, respectively, indicating acceptable model fit. MDD was not included in the CFA or following twin modeling due to the smaller sample size and no significant exposure being identified. Then we used the exposome score to conduct bivariate twin modeling between the exposome score and depressive symptoms. Given the sex differences in prevalence of depressiveness symptoms, the differences in heritability and that sex-limited bivariate models also indicated significant sex differences (Supplementary Table 4) at both age points, we ran the bivariate models separately for males and females.

Figure 3 and Supplementary Table 5 show the path coefficients for the model for exposome score and log-transformed GBI score in young adulthood. Unique environmental factors accounted for 23% and 13% of the covariances in males and females, respectively, while additive genetic factors accounted for 77% in males and 87% in females. In males, standardized variances of E_exposome_ and E_GBI_ were 0.32 (95% CI: 0.26, 0.39) and 0.51 (95% CI: 0.42, 0.62) in males, while the number reduced to 0.25 (95% CI: 0.21, 0.30) and 0.50 (95% CI: 0.42, 0.58) in females, respectively. The remaining share of variance was accounted for by additive genetic effects.

**Figure 3:**
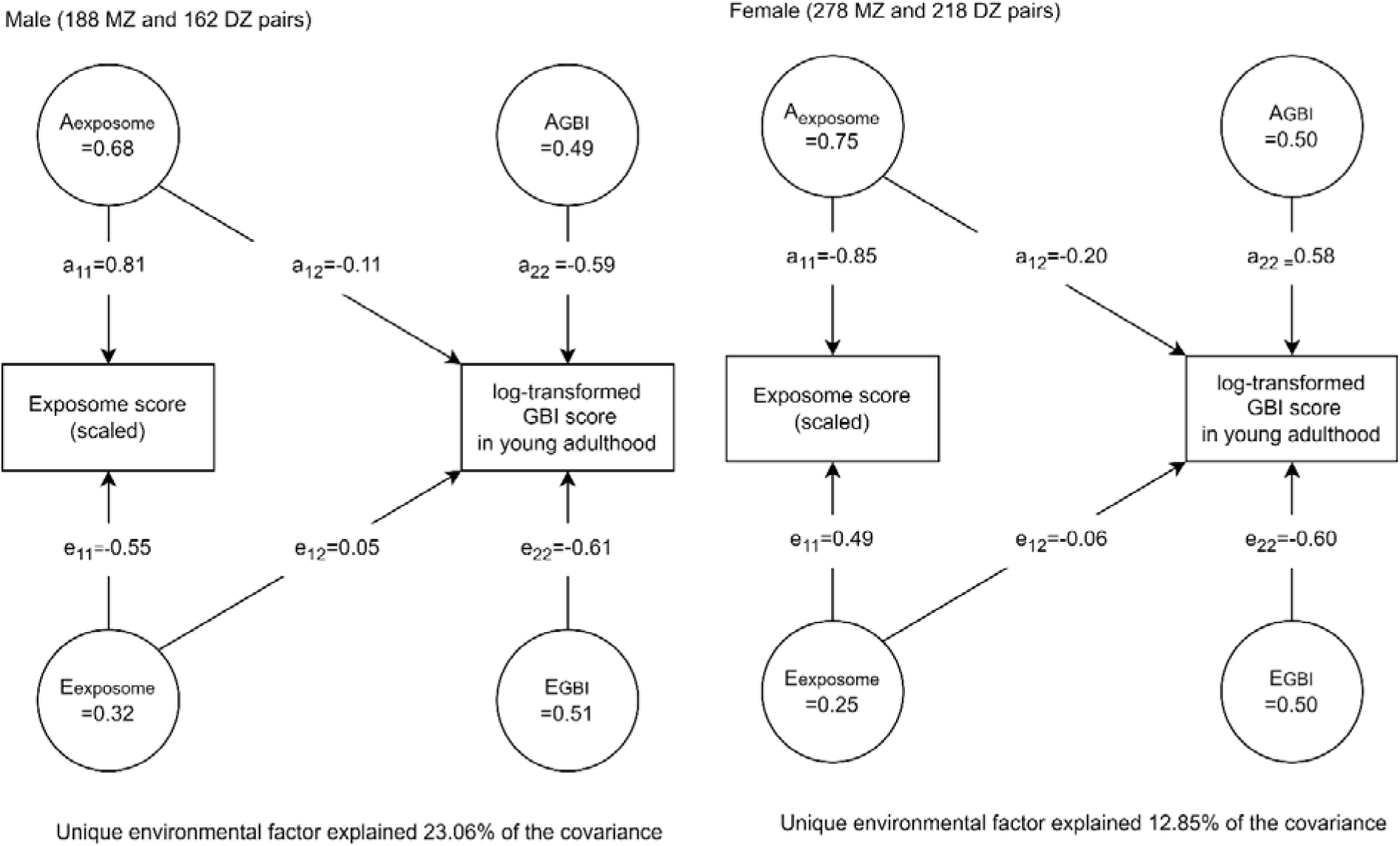
Bivariate Cholesky AE model for the exposome score and log-transformed GBI score in young adulthood ^a^ ^a^ A stands for standardized variance of additive genetic effect. E stands for standardized variance of unique environmental effect. MZ and DZ stand for monozygotic and dizygotic twin pairs, respectively. The 95% confidence intervals of standardized variances and pathway coefficients are presented in Supplementary Table 4.

Supplementary Figure 4 and Supplementary Table 5 show the path coefficients for the model for exposome score and log-transformed GBI score at age 17. Unique environmental factors accounted for 31% and 13% of the covariances in males and females, respectively. Additive genetic factors accounted for 69% in males and 87% in females. The standardized variances of E_exposome_ at age 17 are similar to E_exposome_ in young adulthood regardless of sex. The standardized variance of E_exposome_ is 0.26 (95% CI: 0.22, 0.30) and 0.22 (95% CI: 0.19, 0.25) and of E_GBI_ is 0.64 (95% CI: 0.55, 0.73) and 0.44 (95% CI: 0.38, 0.50) in males and females, respectively. The remaining share of variance was accounted for by additive genetic effects.

Post-hoc mixed model repeated measures (MMRM)

Based on the longitudinal design and 27 significant exposures selected by both ExWASes of log-transformed GBI score, after adjusting for covariates and baseline effect, all the exposures were still significantly associated with log-transformed GBI score in young adulthood. The results are presented in Supplementary Table 6.

## Discussion

Using data on depressive symptoms and diagnosed MDD from the FinnTwin12 study and a wide range of exposures from multiple sources, we applied a two-stage analysis to first screen the exposome and then estimate the environmental sources of correlation between the exposome and depressive symptoms via twin modeling. First, multiple exposures have been identified across domains of family and parents, friend and romantic relationships, school and teachers, and stressful life events, by self-report, which were significantly associated with depressive symptoms in young adulthood and at age 17. In contrast, none of the exposures correlated with the incidence of MDD in young adulthood. Second, after generating an exposome score based on significantly associated exposures, the best-fitting bivariate AE models indicated that unique environmental effects accounted for a marked fraction of the covariance between the exposome score and depressive symptoms. This environmental fraction was higher in males than in females, suggesting a notable sex difference. Our result implies that environmental effects are more impactful compared to genetic effects in males than in females.

Influence from the familial component of the social exposome, especially from the familial atmosphere, was demonstrated by our evidence as having the most substantial impact on depressive symptoms in late adolescence and early adulthood and their trajectory. A large Chinese survey also found that familial factors like cohesion, conflict, and control correlated with the occurrence of depressive symptoms among university students^15^. Other studies have revealed the connection of family triangulation (parent-child coalition and alliance) and satisfaction with depressive symptoms from childhood to late adolescence across countries^16,17^. Fairness (largest protective effect size of GBI at both age points), as a dimension of parentification, was demonstrated as a unique predictor of mental health symptoms^18^. These existing conventional investigations were consistent with ours, while our ExWAS more systematically evaluated a wide range of exposures and reduced the chance of type-I error without any pre-identified hypothesis. Moreover, instead of traditional scales for assessing familial and interpersonal relationships, we treated each scale component as an “independent” exposure in models, which helped us to identify new correlations, detect the relative importance, and prepare for further analysis of more intricate relationships between different components and depressive symptoms.

Results from bivariate twin modeling reveal a complex relationship between genes, environments, and depressive symptoms. Although the unique environmental factor explains a notable amount of covariance between exposome score and depressive symptoms, the additive genetic factor explained relatively more. Many significant exposures were chosen under the guidance of the exposome paradigm, but it does not necessarily imply a pure environmental effect. Many familial influences are considered “inheritable factors” between generations to a certain extent, according to the intergenerational transmission theory. Such effects can be transmitted from parents to children through shared genes but also by shared environments. Early studies have found that life satisfaction or family violence from parents and origin-families led to a significant impact on the development of subsequent similar familial environments among offspring^19,20^. Moreover, we should consider the existence of the gene–environment interaction (G×E), which suggests the different effects of a genotype on disease risk in persons with different environmental exposures^21^. Choi et al.^11^ stratified the ExWAS by polygenic risk scores of major depression and found that some significant factors in the full sample became null in the genetically at-risk sample. Another study suggested the multiple modulation pathways by exposure to DNA methylation, through numerous testing, regarded as the G×E-WAS^22^. Additionally, previous twin studies found geographic confounding in the assessment of A, C, and E variance, possibly attributable to differences in genetic ancestry. Results from the Netherlands Twin Register found 1.8% of the variance in children’s height was captured by regional clustering^23^. In the Netherlands, there were strong genetic differentiations between the north and south, between the east and west, and between the middle band and the rest of the country by PCA on genome-wide data^24^. In the Finnish population, a significant population structure difference is also observed between the east and west parts of country^25^. In brief, the hidden heritable and genetic factors critically influence the association between the exposome and depressive phenotype through various mechanisms, which potentially lead to a propensity to weak associations in our findings.

Furthermore, exposures from the more external domains, particularly in the physical exposome, also showed at most weak connections with depressive symptoms. While it may be the case that the relative importance of the physical exposome is much less than that of the social and familial exposome with respect to depressive symptoms, there are possibly other explanations. First, a more complex structure of the exposome, such as the interaction or correlation between individual exposures and external exposome, may exist. Some previous exposome analyses have indicated this^26,27^, but the ExWAS design cannot characterize it. For instance, the social exposome is an explaining part of the physical exposome, which could not be completely separated. We aim to investigate the complicated effect of the depressive phenotype in the pluralistic platform like machine learning based on our findings in the future. Second, Finland has been ranked very high in the beneficial environmental effect on the child by UNICEF, providing environments with low air pollution, high greenness, safe water, and other constructive aspects relatively equally to most residents in childhood and adolescence^28^. It could explain null results with external living environments due to a lack of individual variation in exposures. Another matter contributing to the large contribution of familial effects is the overlap between interpersonal relationships and depressive symptoms. In a Swedish twin study among females, interpersonal relationships contributed between 18% and 31% of the variance for depressive symptoms^29^. Some personality disorders are tightly connected with interpersonal relationships, for example, borderline, avoidant, and paranoid personality disorder’s liability factors overlapped substantially with MDD’s in particular clusters among Norwegian young adults^30^. This overlapping may have led to an overestimation of the importance of interpersonal relationships.

For social indicators, besides the critical period, various risk models such as accumulation or trajectory may exist, which may also explain the null results. Morrissey and Kinderman confirmed the hypothesis that accumulation of adverse financial hardship negatively affects mental health, but not the hypothesis of critical periods^31^, while our risk model is the “critical period”. Another study demonstrated the complicated effect between changes in racial composition, neighborhood socioeconomic status, and depressive symptoms^32^. The social indicators derived from Statistics Finland’s registers are at the postal code or municipality level, which leads to some concern about the inaccurate measurement of individual’s exposure (information bias).

Several previous ExWAS studies, linking the exposome to mental health, had some similar or heterogeneous results to ours. van de Weijer et al.^10^ identified several social indicators such as safety and income being linked to mental well-being, but the links were weak in our analysis. This may be due to using different outcomes, the older age in their samples, and different statistical methods between the two countries’ authorities^10^. Although Choi et al.’s ExWAS was on the general population in the UK, they also found that a higher frequency of visits with family/friends reduced the odds of depression incidence and Mendelian randomization reinforced the causality of this association^11^. However, we do not have many common variables with Choi et al.^11^ in which they included many lifestyle factors (specific external exposome), while we have more general external exposome variables. Another ExWAS on psychotic experiences identified many stressful life event factors, a result that was similar to our study^8^. Despite the divergent findings, the accumulation of ExWAS findings from different countries, populations, and age groups helps us to enhance our understanding of growing concepts of the exposome on depression, as well as broad mental health. The inclusion of a large number of exposures about interpersonal and person–societal relationships is also an important addition to the existing evidence. Notably, some of the information was provided by the parents, not only the twins. Furthermore, some scientists have raised the concept of an “Eco-Exposome” to thoroughly assess the internal exposome including molecules affected by exogenous exposures^33^, which could be assimilated into further research.

The sex difference is notable. Our previous study found that male twins tend to stay together longer, implying more exposure to any familial impact^34^. In a Swedish study, family structure, conflict, and child disclosure of information to parents were associated with offending behavior in boys, while only one factor was salient in girls^35^. Another British study found that boys in detrimental familial environments were increasingly disadvantaged in school achievement, compared to girls^36^. The evidence hints that males are more easily affected by the family environment, which could explain the higher contribution of E on the covariance between the exposome and depressive symptoms in males. This inference is not certain, and there is contrary evidence^37^. Moreover, sex differences exist in many biological mechanisms regarding how the body neurophysiologically reflects the external environment. Several sex-differentially expressed neurotransmitters or hormones, such as progesterone in females, are involved in systemic dysregulation, inducing depression^38^. Furthermore, environmental endocrine-disrupting chemicals are able to alter neurodevelopment with sex-specific effects at very early developmental stages^39^. In the future, integrating with the internal exposome such as metabolites and other -omics will help us advance the study of sex-difference mechanisms on the relationship between the exposome and depressive phenotype.

As a part of the European Human Exposome Network, our overarching goal is to evaluate the impact of the exposome on human health across various age groups and with respect to multiple outcomes. The present analysis represents one individual analysis, and by pooling our collective efforts important implications for clinical practice can be drawn in the future. Our findings suggest that studies on the familial component of social exposome should be noticed and investigated in the improvement of current therapy. It doesn’t mean that we should ignore the physical exposure group, due to ubiquity, even though their relevance is not salient^40^. In addition, it is imperative to incorporate the consideration of familial effects and genetic liability at the same time for a more thorough understanding in future studies.

There are some other limitations in our study. First, compared to other ExWASes, our sample size is relatively small. Although Chung et al. indicated that a sample size between 1795 and 3625 participants is adequate when using the Bonferroni correction^41^, we did not stratify the ExWAS by sex due to the sample size being reduced by half. Second, we did not further assess the causality. Causal inferences are critical for further policymaking and intervention. Mendelian randomization in larger samples is a future direction. Third, the ExWAS, CFA, and twin modeling were all performed based on the FinnTwin12 cohort, which raises concerns about model overfitting and leakage. Different models with different purposes, hypotheses, and methodologies in two stages reduce the risk of overfitting and leakage. ExWAS was used to identify salient exposure, while CFA and twin modeling were used to explore. The observational unit was each twin-pair in twin modeling, while in ExWAS and CFA it is each individual twin. Replication on other twin cohorts and in family data sets is warranted.

## Conclusion

This study applied a two-stage analysis. First, in ExWAS, we identified that exposures from family and parents, friend and romantic relationships, school and teachers, and stressful life events were significantly associated with depressive symptoms in late adolescence and young adulthood. The family and parent exposures were the most influential. Second, twin modeling between the exposome and depressive symptoms uncovered a complex relationship between genes, environments, and depressive symptoms with sex differences. The findings underline the importance of systematic evaluation of the environmental effects on depressive symptoms and recommend the consideration of genetic effects in future studies.

## Methods

### Study participants

The participants came from the FinnTwin12 cohort, which is a nationwide prospective cohort among all Finnish twins born between 1983 and 1987. First, the overall epidemiological study consisted of all 5184 twins who responded (age 11–12) at wave 1, and there are three general following waves at age 14, 17, and in young adulthood (mean age: 21.9). Moreover, 1035 families with 2070 twins were invited to take part in an intensive study with psychiatric interviews, some biological samples, and additional questionnaires^42^. At age 14 (wave 2), 1854 twins participated.

They were then invited to participate again as young adults (wave 4) of the study. Psychiatric interviews in young adulthood were completed for 1347 twins in the intensive study, including assessment of MDD using the Semi-Structured Assessment for Genetics of Alcohol based on Diagnostic and Statistical Manual of Mental Disorders IV criteria^43,44^. The twins also completed questionnaires on health, health behaviors, work, and multiple psychological scales. An updated review has been published^45^.

### Measures

The primary outcome is the short-version GBI scores in young adulthood. It is a self-reported inventory to evaluate the occurrence of depressive symptoms, which is composed of 10 Likert-scale questions^46^. The total score ranges from 0 to 30, and a higher score implies more depressive symptoms occurred. There are two secondary outcomes: GBI scores at age 17 and incidence of MDD in young adulthood.

In total, we curated 385 environmental exposures under the concept of the Equal-Life project^47^ from multiple sources and group them into 12 domains. Air pollution exposures came from the annual average air quality of each observation station from the Finnish Meteorological Institute. Domains of building, blue and green spaces, population density, and a part of geocoordinates were from Equal-Life enrichment. Their description can be found in a previous study^48^ and is presented in Supplementary Note 1. Exposures from prenatal exposures, passive smoking, family and parents, friend and romantic relationships, school and teachers, and stressful life events domains were from FinnTwin12 questionnaires by self-report or parent-report and are described in a published review^45^. Social indicators were from Statistics Finland and described in Supplementary Note 1. Except for FinnTwin12 questionnaires, exposures from other sources were linked to individual twins via EUREF-FIN geocoordinates. The full residential history from birth onward until 2020 of the twins was obtained as geocoordinates and dates of moving in and out of specific addresses from the Digital and Population Data Services Agency in Finland^34^. The types of exposures are continuous, binary, and categorical. Considering the temporality, we included repeated exposures for the “critical period” risk model, and Supplementary Figure 5 presents the timeline of the study. There are three exposure inclusion criteria: 1) twins have available residential history; 2) twins and their family completed at least one questionnaire at any wave; and 3) the percentage of missing values is less than 20% in ExWAS. The codenames of each exposure were developed from the description as closely as possible, and their domains, resources, and dates were presented in Supplementary Table 1.

For analysis of outcomes in young adulthood, we *a priori* identified seven covariates: sex (male, female), zygosity (monozygotic (MZ), DZ, unknown), parental education (limited, intermediate, high)^49^, smoking (never, former, occasional, current), work status (full-time, part-time, irregular, not working), secondary level school (vocational, senior high school, none), and age. The latter four variables were reported by twins as young adults (wave 4). For analysis of outcome at age 17, sex, zygosity, parental education, smoking (reported at age 17) remained. Study and working status (neither study nor work, only study, only work) were included when most participants were in school at age 17 (wave 3). The inclusion of covariates, besides sex, zygosity, and age, was based on the previous literature, which shows correlations with the environment and depressive symptoms^50–52^. Parental education was adjusted for to represent the family resources and resilience^49^.

### Data pre-processing and descriptive statistics

Participants missing information on outcome or covariates were excluded from the corresponding age’s analyses. Due to the skewness of the GBI score, we added one to the GBI score and log-transformed it. Appropriate regrouping was conducted for categorical exposures, and then we used multivariate imputation by chained equation to replace the missing values of exposures. As a dimension reduction technique, PCA was utilized to measure the proportion of total variability of all included exposures attributed to each PC and visually assess the potential clusters of exposures (correlated) based on the two-dimensional coordinate with the first and second components. It was only conducted for outcomes of GBI at age 17 and in young adulthood, not for the incidence of MDD.

### Exposome-wide association study

To conduct the ExWAS, a generalized linear regression model with Gaussian distribution (essentially linear regression) for the outcomes of log-transformed GBI score was repeatedly performed for each exposure. We used Bonferroni correction by the number of effective tests (calculated by PCA) to adjust for multiple testing and account for correlation between exposures^53^. Covariates were adjusted and the cluster effect of sampling based on families of twin pair was controlled for by the robust standard error. For the outcome of the incidence of MDD, the distribution was switched to be binomial. The number of included exposures of secondary outcomes was smaller due to the third exposure inclusion criteria and the sample size varied, thus the P value thresholds varied. Due to categorical exposures, the number of P values was higher than the number of exposures. The R package “rexposome” was used^54^.

### Generating exposome score

Based on the significant exposures selected from the ExWAS, confirmatory factor analysis was used to estimate an exposome score, preparing for the following twin modeling. According to the concept of the environment’s totality, we indicated a one-factor structure for the exposome. CFA assumes the correlation between exposures due to the exposome score and verifies it based on structural equation modeling as theory-driven. We used maximum likelihood to estimate the score and SRMR to evaluate the model fit^55^. The cluster effect was controlled like before. Due to multiple subgroups in categorical exposures, we included the whole exposure variable when there was at least one subgroup that was significant compared to the reference in ExWAS. The Stata package “sem” was used. The coefficients of significant exposures were presented in Supplementary Tables 7 and 8 for outcomes of GBI in young adulthood and at age 17, respectively. Additionally, we also conducted exploratory factor analysis (EFA) estimated by maximum likelihood with 100 optimizations, whereas a large number of retained factors indicated potential overfitting of EFA.

### Twin modeling

In twin modeling, the genetic effect is usually divided into additive and dominant genetic effects^56^. Since MZ twins are roughly genetically identical and DZ twins share roughly half of their segregating genes, the correlation of A is set to 1.0 and 0.5 and of D is set to 1.0 and 0.25 within MZ and DZ twin pairs, respectively. The epistatic effect is a part of A. The environmental effect is also divided into two components: common environment whose correlation is assumed to be 1.0 regardless of zygosity, and unique environment (no correlation), which includes unmeasured errors. The use of the twin model assumes the absence of assortative mating for the trait under study among the parents and equal effects of the environment by zygosity.

The intrapair correlations of GBI in DZ (ρ=0.22 in young adulthood and =0.16 at age 17) and MZ (ρ=0.52 in young adulthood and =0.51 at age 17) indicated to use an ADE model initially, instead of the ACE model (ρ_MZ_>2ρ_DZ_). Due to only using the twin pair design, instead of the extended family design, we could not use an ACDE model. The saturated twin model was performed to test the assumptions of equal means and variances for twin order and for zygosity, via constraint means and variances, and to detect the sex difference via sex limitation. In the saturated model (Supplementary Table 9), the AIC and likelihood ratio test between models suggested that the assumptions were basically met. Results of the sex-limitation saturated model (Supplementary Table 9) indicated a significant sex difference.

Finally, to assess how the current exposome score explains the variance of depressive symptoms, we employed the bivariate Cholesky AE model to fit the exposome score and log-transformed GBI score (Supplementary Figure 6) at both age points, which efficiently decomposes the phenotypic correlation and offers the attribution (%) to genetic and environmental factors^57^. Two latent factors (A_exposome_ and E_exposome_) influence both the exposome score (a_11_ and e_11_) and log-transformed GBI score (a_21_ and e_21_), and another two latent factors (A_GBI_ and E_GBI_) only influence the log-transformed GBI score (a_22_ and e_22_). The overall correlation between the exposome score and GBI could be calculated as a_11_ * a_12_ + e_11_ * e_12_. Variances of A_exposome_, E_exposome,_ A_GBI_, and E_GBI_ were calculated as 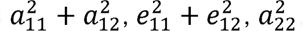, and 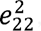, respectively. We also re-assess the sex difference via an additional sex-limited saturated bivariate twin model.

Only full MZ and DZ twin pairs were included in the twin modeling. We dropped the opposite-sex DZ pairs and stratified the univariate and bivariate twin models by sex. The characteristics of included and excluded individual twins in the twin modeling are presented in Supplementary Table 10, and we did not observe a large difference suggesting low selection bias risk due to sex, zygosity, and twin pair. Age, reported in the young adulthood survey, was adjusted in univariate and bivariate models for the outcome in young adulthood. The R package “OpenMx” was used^58^.

Post-hoc mixed models for repeated measures

Based on the exposures significantly associated with GBI at both time points, we performed the MMRM, as a post-hoc analysis, to further explore the effects on the trajectory of depressive symptoms. This method analyzes the influence on the log-transformed GBI in young adulthood by both exposures of interest (fixed effect) and “baseline” log-transformed GBI at age 17 (random effect)^59^. The sample size and covariates of the MMRM were the same as the ExWAS of log-transformed GBI score in young adulthood. The cluster effect was controlled by the robust standard error. The multiple testing was controlled by the false discovery rate (Q value <0.05 was considered statistically significant).

## List of abbreviations

AIC: Akaike information criterion
CFA: Confirmatory factor analysis
DZ: Dizygotic
EFA: Exploratory factor analysis
ExWAS: Exposome-wide association study
GBI: General behavior inventory
G×E: Gene–environment interaction
MDD: Major depressive disorder
MMRM: Mixed model for repeated measures
MZ: Monozygotic
PCA: Principal component analysis
SRMR: Standardized root mean square residual

## Data availability

The FinnTwin12 data is not publicly available due to the restrictions of informed consent. However, the FinnTwin12 data is available through the Institute for Molecular Medicine Finland (FIMM) Data Access Committee (DAC) for authorized researchers who have IRB/ethics approval and an institutionally approved study plan. To ensure the protection of privacy and compliance with national data protection legislation, a data use/transfer agreement is needed, the content and specific clauses of which will depend on the nature of the requested data.

## Code availability

No new software, package, and algorithm were developed. All code for data cleaning and analysis associated with the current submission is available upon reasonable request to the corresponding author.

## Acknowledgement

We would like to acknowledge Dr. Anttila Pia and Dr. Hellén Heidi from the Finnish Meteorological Institute for providing the annual summary of air quality data in Finland. We would like to appreciate Albert Ambros and Bruno Raimbault from ISGlobal for the calculation of the exposome for this study. We would like to thank Xavier Escribà-Montagut from ISGlobal for technical support on the R package “rexposome”. FinnTwin12 wishes to thank all participating twins, their parents, and teachers.

## Inclusion & Ethics

The ethics committee of the Department of Public Health of the University of Helsinki (Helsinki, Finland), the ethics committee of the Helsinki University Central Hospital District (Helsinki, Finland), and the Institutional Review Board of Indiana University (Bloomington, Indiana, USA) approved the FinnTwin12 study protocol. All participants and their parents/legal guardians gave informed written consent to participate in the study. The authors assert that all procedures contributing to this work comply with the ethical standards of the relevant national and institutional committees on human experimentation and with the Helsinki Declaration of 1975, as revised in 2008.

## Reference

1. GBD 2019 Mental Disorders Collaborators. Global, regional, and national burden of 12 mental disorders in 204 countries and territories, 1990-2019: a systematic analysis for the Global Burden of Disease Study 2019. The lancet. Psychiatry 9, 137–150 (2022).

2. Shorey, S., Ng, E. D. & Wong, C. H. J. Global prevalence of depression and elevated depressive symptoms among adolescents: A systematic review and meta-analysis. Br. J. Clin. Psychol. 61, 287–305 (2022).

3. Köhler, C. A. et al. Mapping risk factors for depression across the lifespan: An umbrella review of evidence from meta-analyses and Mendelian randomization studies. J. Psychiatr. Res. 103, 189–207 (2018).

4. Mair, C., Roux, A. V. D. & Galea, S. Are neighbourhood characteristics associated with depressive symptoms? A review of evidence. J. Epidemiol. Community Health 62, 940 LP – 946 (2008).

5. Wild, C. P. Complementing the Genome with an “Exposome”: The Outstanding Challenge of Environmental Exposure Measurement in Molecular Epidemiology. Cancer Epidemiol. Biomarkers Prev. 14, 1847–1850 (2005).

6. Guloksuz, S., van Os, J. & Rutten, B. P. F. The Exposome Paradigm and the Complexities of Environmental Research in Psychiatry. JAMA Psychiatry 75, 985–986 (2018).

7. Zheng, Y. et al. Design and methodology challenges of environment-wide association studies: A systematic review. Environ. Res. 183, 109275 (2020).

8. Lin, B. D. et al. Nongenetic Factors Associated With Psychotic Experiences Among UK Biobank Participants: Exposome-Wide Analysis and Mendelian Randomization Analysis. JAMA Psychiatry 79, 857–868 (2022).

9. Ni, M. Y. et al. Determinants of physical, mental and social well-being: a longitudinal environment-wide association study. Int. J. Epidemiol. 49, 380–389 (2020).

10. van de Weijer, M. P. et al. Expanding the environmental scope: an environment-wide association study for mental well-being. J. Expo. Sci. Environ. Epidemiol. 32, 195–204 (2022).

11. Choi, K. W. et al. An Exposure-Wide and Mendelian Randomization Approach to Identifying Modifiable Factors for the Prevention of Depression. Am. J. Psychiatry 177, 944– 954 (2020).

12. Leffers, H. C. B., Lange, T., Collins, C., Ulff-Møller, C. J. & Jacobsen, S. The study of interactions between genome and exposome in the development of systemic lupus erythematosus. Autoimmun. Rev. 18, 382–392 (2019).

13. Wichers, M. et al. Mechanisms of gene–environment interactions in depression: evidence that genes potentiate multiple sources of adversity. Psychol. Med. 39, 1077–1086 (2009).

14. Medda, E. et al. The response to oxidative stress and metallomics analysis in a twin study: The role of the environment. Free Radic. Biol. Med. 97, 236–243 (2016).

15. Yu, Y., et al. The Role of Family Environment in Depressive Symptoms among University Students: A Large Sample Survey in China. PLoS One 10, e0143612 (2015).

16. Stavropoulos, V., Lazaratou, H., Marini, E. & Dikeos, D. Low Family Satisfaction and Depression in Adolescence: The Role of Self-Esteem. J. Educ. Dev. Psychol. 5, 109–118 (2015).

17. Wang, L. & Crane, D. R. The Relationship Between Marital Satisfaction, Marital Stability, Nuclear Family Triangulation, and Childhood Depression. Am. J. Fam. Ther. 29, 337–347 (2001).

18. Hooper, L. M. & Wallace, S. A. Evaluating the Parentification Questionnaire: Psychometric Properties and Psychopathology Correlates. Contemp. Fam. Ther. 32, 52–68 (2010).

19. Cappell, C. & Heiner, R. B. The intergenerational transmission of family aggression. J. Fam. Violence 5, 135–152 (1990).

20. Powdthavee, N. & Vignoles, A. Mental Health of Parents and Life Satisfaction of Children: A Within-Family Analysis of Intergenerational Transmission of Well-Being. Soc. Indic. Res. 88, 397–422 (2008).

21. Ottman, R. Gene–Environment Interaction: Definitions and Study Design. Prev. Med. (Baltim). 25, 764–770 (1996).

22. Carreras-Gallo, N. et al. The early-life exposome modulates the effect of polymorphic inversions on DNA methylation. Commun. Biol. 5, 455 (2022).

23. Tamimy, Z. et al. Multilevel Twin Models: Geographical Region as a Third Level Variable. Behav. Genet. 51, 319–330 (2021).

24. Abdellaoui, A. et al. Population structure, migration, and diversifying selection in the Netherlands. Eur. J. Hum. Genet. 21, 1277–1285 (2013).

25. Kerminen, S. et al. Fine-Scale Genetic Structure in Finland. G3 Genes|Genomes|Genetics 7, 3459–3468 (2017).

26. Ohanyan, H. et al. Machine learning approaches to characterize the obesogenic urban exposome. Environ. Int. 158, 107015 (2022).

27. Ohanyan, H. et al. Associations between the urban exposome and type 2 diabetes: Results from penalised regression by least absolute shrinkage and selection operator and random forest models. Environ. Int. 170, 107592 (2022).

28. UNICEF Office of Research. Places and Spaces Environments and children’s well-being. (2022).

29. Spotts, E. L. et al. Accounting for depressive symptoms in women: a twin study of associations with interpersonal relationships. J. Affect. Disord. 82, 101–111 (2004).

30. Reichborn-Kjennerud, T. et al. Major depression and dimensional representations of DSM-IV personality disorders: a population-based twin study. Psychol. Med. 40, 1475–1484 (2010).

31. Morrissey, K. & Kinderman, P. The impact of childhood socioeconomic status on depression and anxiety in adult life: Testing the accumulation, critical period and social mobility hypotheses. SSM - Popul. Heal. 11, 100576 (2020).

32. Lee, H. & Estrada-Martínez, L. M. Trajectories of Depressive Symptoms and Neighborhood Changes from Adolescence to Adulthood: Latent Class Growth Analysis and Multilevel Growth Curve Models. Int. J. Environ. Res. Public Health 17, (2020).

33. Scholz, S. et al. The Eco-Exposome Concept: Supporting an Integrated Assessment of Mixtures of Environmental Chemicals. Environ. Toxicol. Chem. 41, 30–45 (2022).

34. Wang, Z., Whipp, A., Heinonen-Guzejev, M. & Kaprio, J. Age at Separation of Twin Pairs in the FinnTwin12 Study. Twin Res. Hum. Genet. 1–7 (2022) doi:DOI: 10.1017/thg.2022.17.

35. Nilsson, E.-L. Analyzing Gender Differences in the Relationship between Family Influences and Adolescent Offending among Boys and Girls. Child Indic. Res. 10, 1079–1094 (2017).

36. Mensah, F. K. & Kiernan, K. E. Gender differences in educational attainment: Influences of the family environment. Br. Educ. Res. J. 36, 239–260 (2010).

37. Blaauboer, M. & Mulder, C. H. Gender differences in the impact of family background on leaving the parental home. J. Hous. Built Environ. 25, 53–71 (2010).

38. Sikes-Keilp, C. & Rubinow, D. R. In search of sex-related mediators of affective illness. Biol. Sex Differ. 12, 55 (2021).

39. Rebuli, M. E. & Patisaul, H. B. Assessment of sex specific endocrine disrupting effects in the prenatal and pre-pubertal rodent brain. J. Steroid Biochem. Mol. Biol. 160, 148–159 (2016).

40. Maria, F. et al. Association of Long-Term Exposure to Traffic-Related Air Pollution with Blood Pressure and Hypertension in an Adult Population–Based Cohort in Spain (the REGICOR Study). Environ. Health Perspect. 122, 404–411 (2014).

41. Chung, M. K., Buck Louis, G. M., Kannan, K. & Patel, C. J. Exposome-wide association study of semen quality: Systematic discovery of endocrine disrupting chemical biomarkers in fertility require large sample sizes. Environ. Int. 125, 505–514 (2019).

42. Whipp, A. M. et al. Early adolescent aggression predicts antisocial personality disorder in young adults: a population-based study. Eur. Child Adolesc. Psychiatry 28, 341–350 (2019).

43. Bucholz, K. K. et al. A new, semi-structured psychiatric interview for use in genetic linkage studies: a report on the reliability of the SSAGA. J. Stud. Alcohol 55, 149–158 (1994).

44. Bell, C. C. DSM-IV: Diagnostic and Statistical Manual of Mental Disorders. JAMA 272, 828–829 (1994).

45. Rose, R. J. et al. FinnTwin12 Cohort: An Updated Review. Twin Res. Hum. Genet. 22, 302– 311 (2019).

46. Kokko, K. & Pulkkinen, L. Unemployment and Psychological Distress: Mediator Effects. J. Adult Dev. 5, 205–217 (1998).

47. van Kamp, I. et al. Early environmental quality and life-course mental health effects: The Equal-Life project. Environ. Epidemiol. 6, (2022).

48. Julvez, J. et al. Early life multiple exposures and child cognitive function: A multi-centric birth cohort study in six European countries. Environ. Pollut. 284, 117404 (2021).

49. Huppertz, C. et al. The effects of parental education on exercise behavior in childhood and youth: a study in Dutch and Finnish twins. Scand. J. Med. Sci. Sports 27, 1143–1156 (2017).

50. Salmela-Aro, K. et al. Depressive Symptoms and Career-Related Goal Appraisals: Genetic and Environmental Correlations and Interactions. Twin Res. Hum. Genet. 17, 236–243 (2014).

51. Edelbrock, C., Rende, R., Plomin, R. & Thompson, L. A. A Twin Study of Competence and Problem Behavior in Childhood and Early Adolescence. J. Child Psychol. Psychiatry 36, 775–785 (1995).

52. Richard J. Rose, J. K. Genes, Environments, and Adolescent Substance Use: Retrospect and Prospect from the FinnTwin Studies. Acta Psychologica Sinica vol. 40 1062–1072 (2008).

53. Li, M.-X., Yeung, J. M. Y., Cherny, S. S. & Sham, P. C. Evaluating the effective numbers of independent tests and significant p-value thresholds in commercial genotyping arrays and public imputation reference datasets. Hum. Genet. 131, 747–756 (2012).

54. Hernandez-Ferrer, C. et al. Comprehensive study of the exposome and omic data using rexposome Bioconductor Packages. Bioinformatics 35, (2019).

55. Kline, R. B. Principles and Practice of Structural Equation Modeling, Fourth Edition. (Guilford Publications, 2015).

56. Polderman, T. J. C. et al. Meta-analysis of the heritability of human traits based on fifty years of twin studies. Nat. Genet. 47, 702–709 (2015).

57. Røysamb, E. & Tambs, K. The beauty, logic and limitations of twin studies. Nor. Epidemiol. 26, (2016).

58. Neale, M. C. et al. OpenMx 2.0: Extended Structural Equation and Statistical Modeling. Psychometrika 81, 535–549 (2016).

59. Detry, M. A. & Ma, Y. Analyzing Repeated Measurements Using Mixed Models. JAMA 315, 407–408 (2016).

