## Supplementary Table 1-10 for "Systematic evaluation of the environmental effect on depressive symptoms in late adolescence and early adulthood: exposome-wide association study and twin modeling"

**Zhiyang Wang et al. Supplemental material 2**

**Content of supplemental material 2**

Supplementary Note 1: Additional description of exposures

Supplementary Figure 1: Principal component analysis for exposures in young adulthood (A) and at age 17 (B)

Supplementary Figure 2: Association results between exposure and incidence of MDD, adjusted for covariates ^a^

Supplementary Figure 3: Association results between exposure and log-transformed GBI score at age 17, adjusted for covariates ^a^

Supplementary Figure 4: Bivariate Cholesky AE model for the exposome score and log-transformed GBI score at age 17 ^a^

Supplementary Figure 5: Calendar timeline of included exposures and outcomes

Supplementary Figure 6: Diagram of bivariate Cholesky AE decomposition model (r: correlation)

Supplementary Reference

Supplementary Note 1: Additional description of exposures

This study is a part of the EU project Equal-life (www.equal-life.eu). The project integrates knowledge and sources from different disciplines and institutions across Europe to develop and utilize the exposome concept and investigate the relationship between the exposome and mental health in children and adolescents. The introduction paper has been published^1^. A part of included exposures was produced from the Equal-life project: blue and green spaces, building, population density, and a part of geocoordinate domains.

Exposures of blue and green spaces domain provide information about the distance, type, and size of the closer green and blue spaces. The EU defines this as living within 300 m of a public open area with more than 5000 m^2^ (Europe 2003) (or within a 15-minute walk). The Europe-wide Urban Atlas (UA)^2^ and CORINE Land Cover^3^ were used to extract maps of urban and natural green and blue spaces. We decided to use these two datasets due to the highest resolution of UA and high coverage of CORINE with the scarce resolution. There are another two indices: Normalized Difference Vegetation Index (NDVI) and Modified Soil Adjusted Vegetation Index (MSAVI) to characterize the vegetation density within buffer areas of 100, 300, and 500 m radii around the geocodes. NDVI quantifies vegetation greenness and is useful in understanding vegetation density and assessing changes in plant health^4^. It is calculated as a ratio between the red (R) and near-infrared (NIR) values in a traditional fashion^4^. MSAVI minimizes the effect of bare soil on the Soil Adjusted Vegetation Index, which corrects NDVI for the influence of soil brightness in areas where vegetative cover is low^5^. It is calculated as a ratio between the R and NIR values with an inductive function applied to maximize the reduction of soil effects on the vegetation signal^5^. Their source is the U.S. Geological Survey Landsat data via Google Earth Engine. Moreover, tree cover indicators estimate the percentage of horizontal ground in the 3 buffer areas at 30 m resolution covered by woody vegetation greater than 5 meters in height, and they are from the Global Forest Cover Change data products through NASA^6^.

Exposures of building domain include “building density”, which is the percentage of area covered by buildings in the three buffer areas and the percentage of impervious area capturing the percentage of soil sealing. Impervious areas are characterized by the substitution of the original (semi-) natural land cover or water surface with an artificial, often impervious cover. These indicators are derived from different data sources. Global Human Settlement (GHS) is for “building density”^7^ and Impervious Surface Area (ISA) is for “percentage of impervious area”^8^, covering different time periods.

Population density is counted within the three buffer areas around the geocode, derived as the weighted sum of the population count indicated in each pixel where the buffer area intersects the population raster data. Worldpop’s (WP) estimation models have been developed, refined, and implemented to produce global multi-temporal 100x100 m datasets for each year^9^. The assumption is that no settlement dataset is accurate enough to identify all residential settlements/buildings globally. The modeling is unconstrained, making predictions about population numbers for all 100x100 m of land grid cells globally for each year through disaggregating a census database. We also consider the GHS population data^7^. The temporal resolution of GHS is good as it goes back to 1975, but these models are only available at two time points in this study: 2000 and 1990^7^. Spatial resolution is available at two levels: 250 m and 1000 m.

A part of geocoordinates exposures, from Equal-life, is the elevation and slope within three buffer areas around the geocode. They are derived from the EU-DEM (digital elevation model) with overall high fundamental vertical accuracy^10^. It ensures that water features are adequately represented and consistent with the hydrography layer^10^. In areas above 60 degrees north, the EU-DEM generation process is supported by other DEM data sources provided by the Joint Research Center’s Global Surface Water dataset. Water features are flattened (oceans, lakes) and stepped (rivers) based on the hydrography data^10^.

Another large part of exposure, social indicators, came from Statistics Finland, which is the national statistical institution in Finland. Generally, we included 21 social indicators, and some indicators contain more than one variable. The statistical reference time point for the population and households is the latest day or week of the statistical year. Most of the variables are presented at the postal code level (2022 version), while the offenses and voting indicators are presented at the municipality level (2022 version). Then, we used the map, containing 2022’s postal code and municipality information, provided by Esri Finland^11^, to link the twins and social indicators in 1990, 2000, and 2006. Some variables are only available after 1995. Time points of voting indicators are slightly different due to dates of elections in Finland.

Supplementary Figure 1: Principal component analysis for exposures in young adulthood (A, individual twin n=3025) and at age 17 (B, individual twin n=1236)

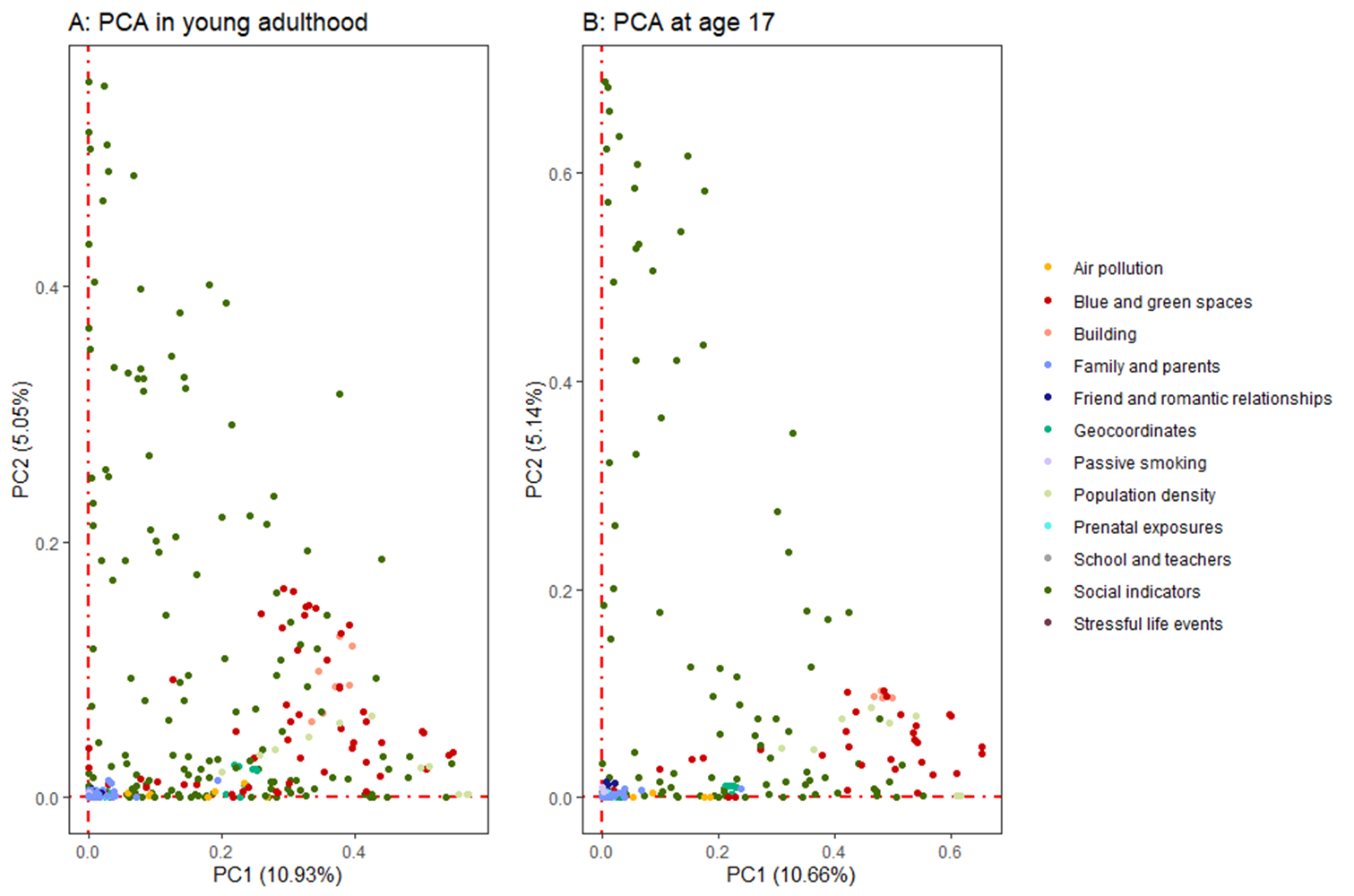

Supplementary Figure 2: Association results between exposure and incidence of MDD, adjusted for covariates (individual twin n=1236) , using generalized binomial regression ^a^

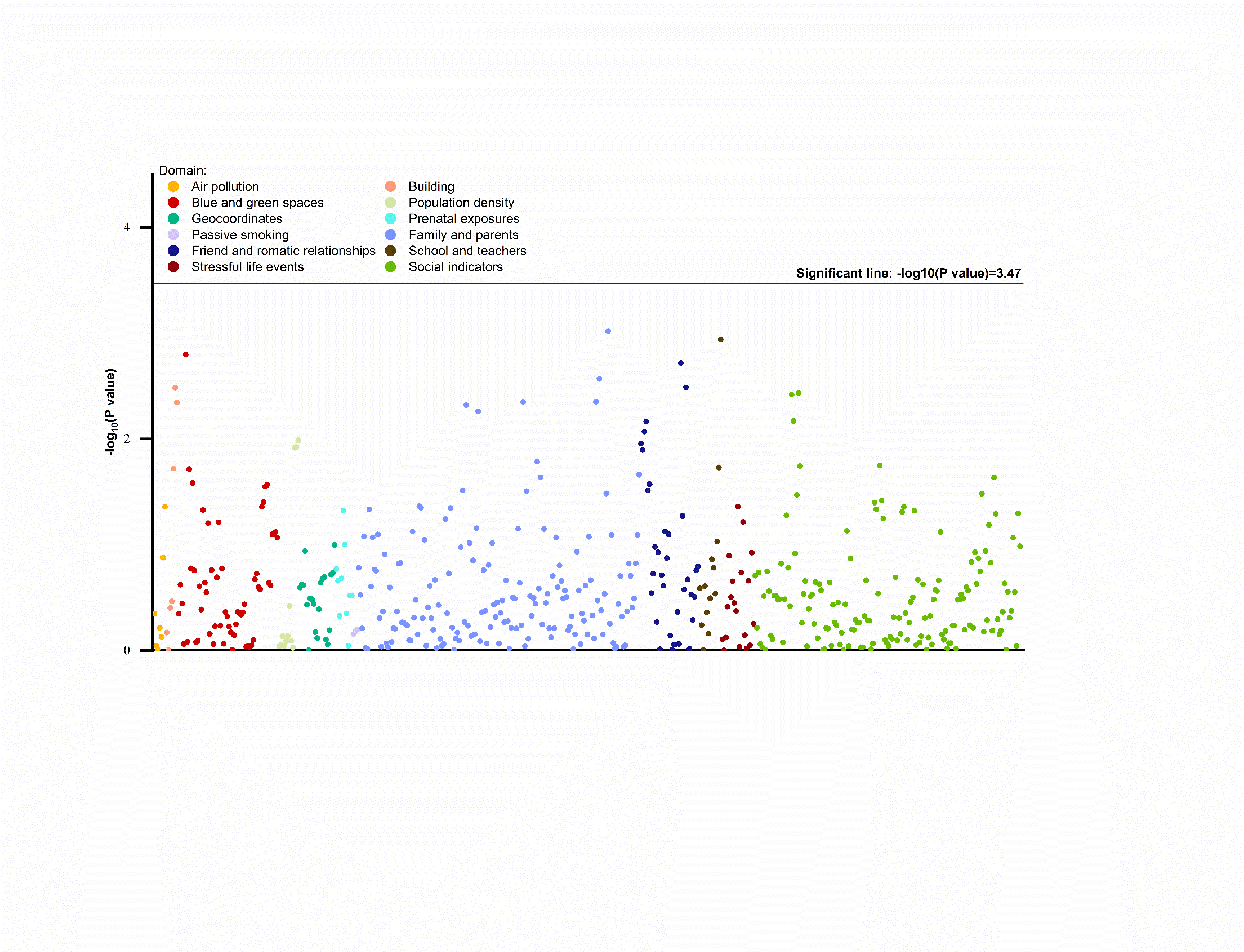

^a^ The adjusted covariates were: sex, zygosity, parental education, smoking in young adulthood, work status in young adulthood, secondary level school in young adulthood, and age when twins provided the GBI assessment in young adulthood

Supplementary Figure 3: Association results between exposure and log-transformed GBI score at age 17, adjusted for covariates (individual twin n=4127), using generalized linear regression ^a^

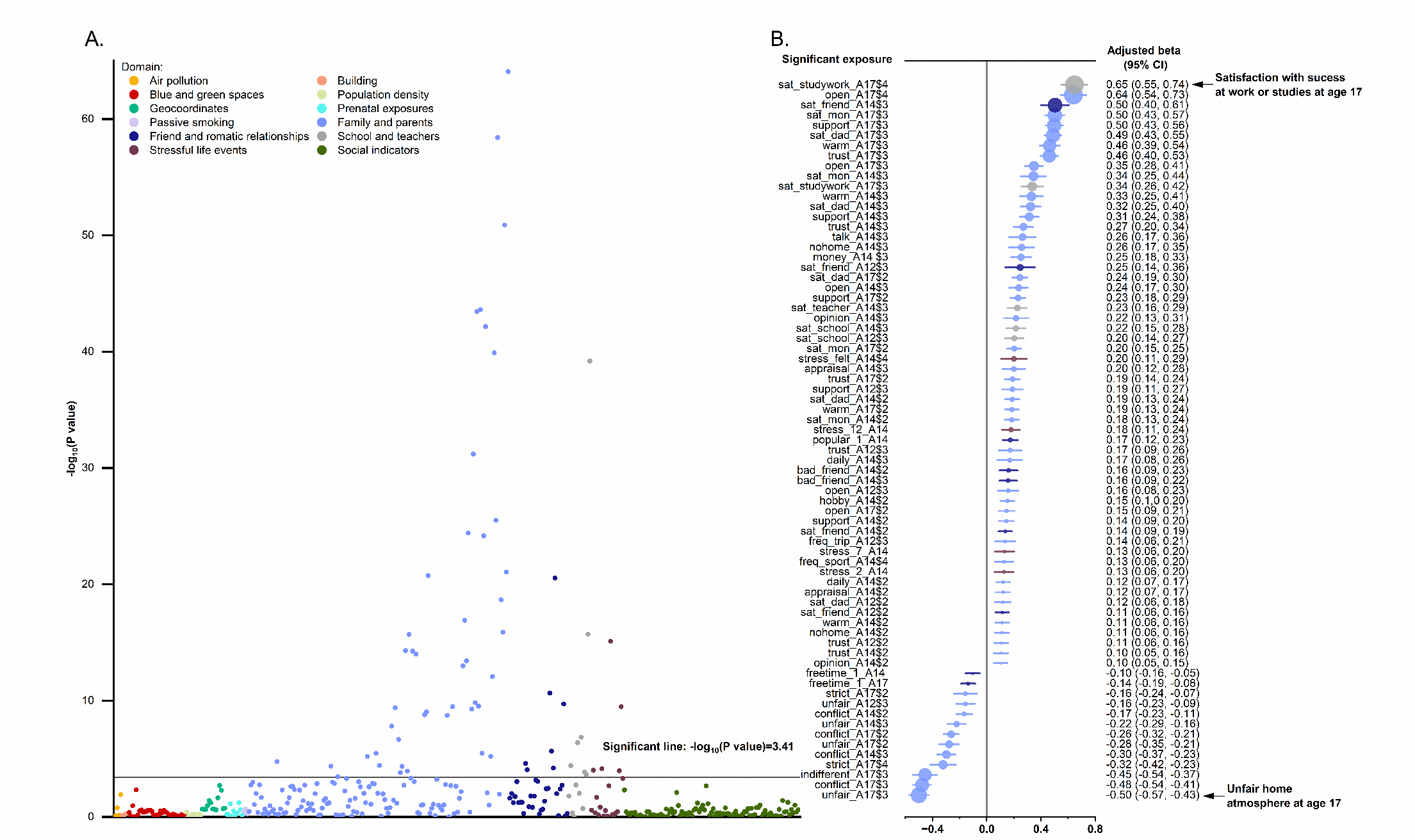

^a^ Panel A is a Manhattan association plot for exposures in relation to log-transformed GBI score at age 17. The y-axis is showing statistical significance as –log10(P value) for the adjustment for multiple testing. Panel B presents the adjusted beta for significant exposures in descending order (from harmful to protective). In panel B, the center dot and bar present the effect size (coefficient of linear regression) and 95% confidence interval, and the size of the dots presents the effect size relatively. The color legend applies to both Panel A (Manhattan association plot) and B (forest plot). The adjusted covariates were: sex, zygosity, parental education, smoking at age 17, and study and working status at age 17.

Supplementary Figure 4: Bivariate Cholesky AE model for the exposome score and log-transformed GBI score at age 17 (twin pair n=1000)^a^

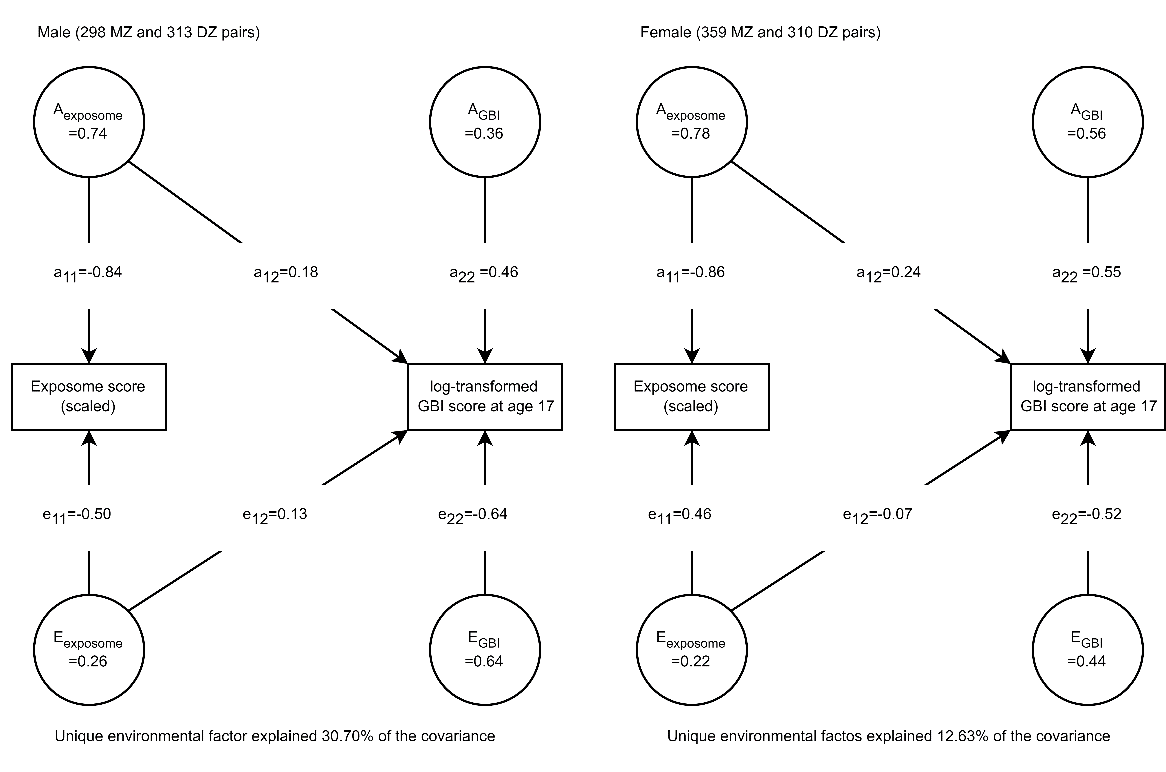

^a^ A stands for standardized variance of additive genetic effect. E stands for standardized variance of unique environmental effect. MZ and DZ stand for monozygotic and dizygotic twin pairs, respectively. The 95% confidence intervals of standardized variances and pathway coefficients are presented in Supplementary Table 4.

Supplementary Figure 5: Calendar timeline of included exposures and outcomes

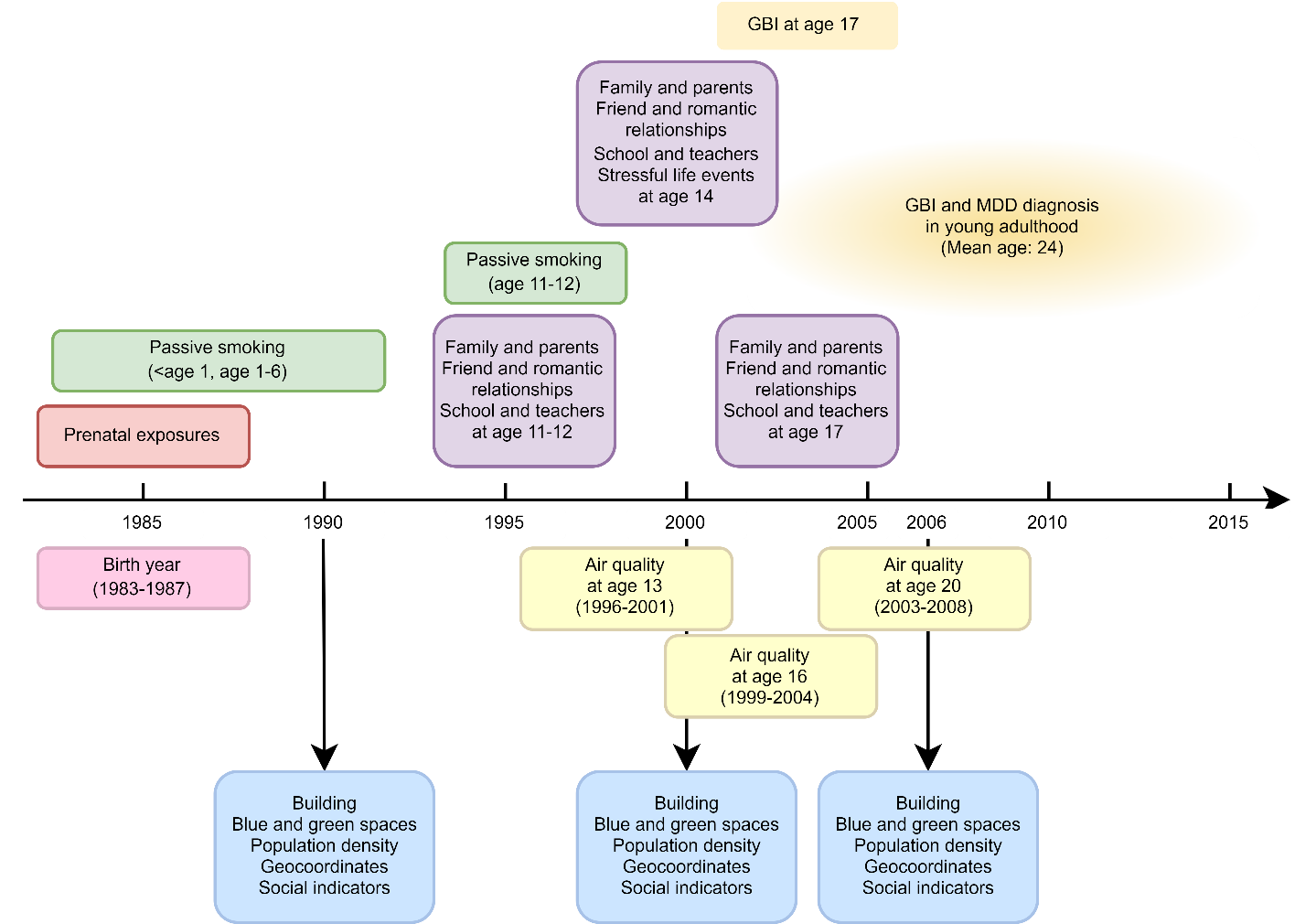

Supplementary Figure 6: Diagram of bivariate Cholesky AE decomposition model (r: correlation)

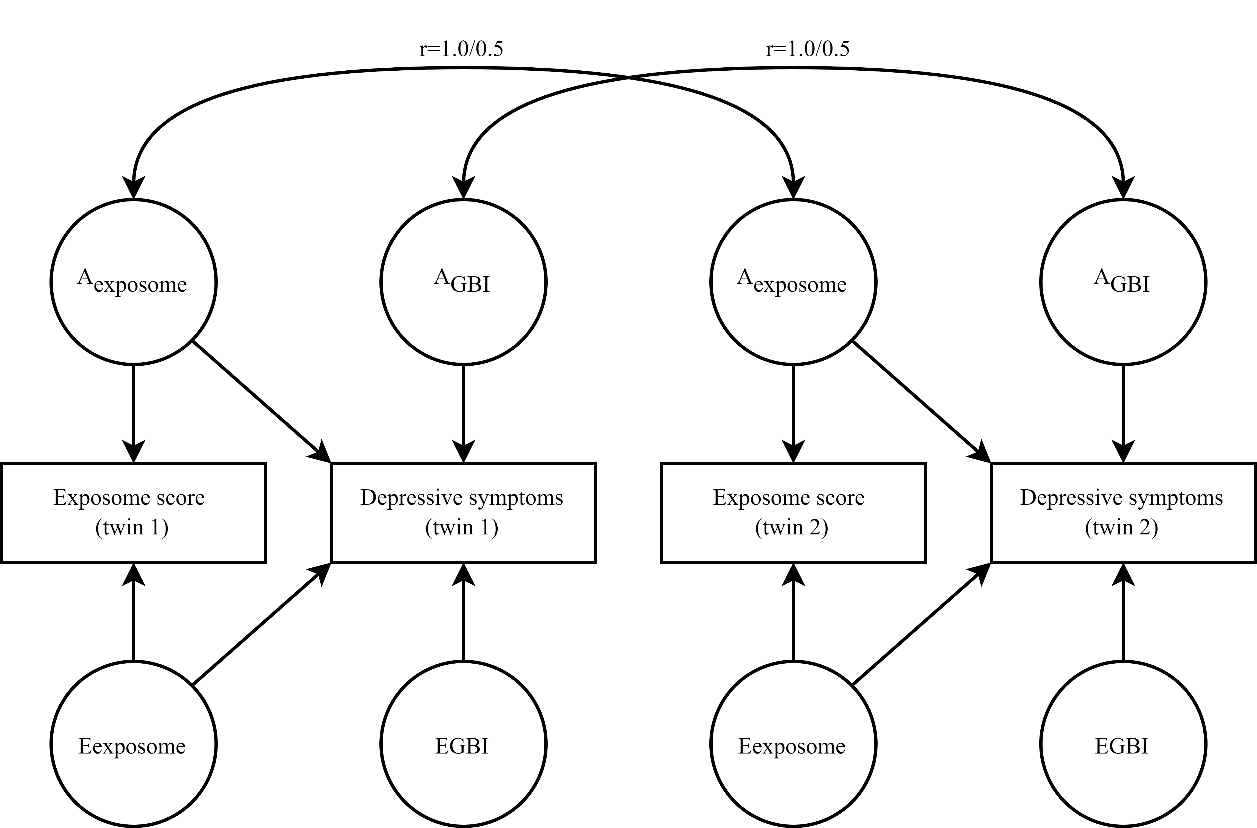

Supplementary Reference

1.	van Kamp, I. *et al.* Early environmental quality and life-course mental health effects: The Equal-Life project. *Environ. Epidemiol.* **6**, (2022).

2.	Urban Atlas LCLU 2012. http://land.copernicus.eu/local/urban-atlas/urban-atlas-2012/view (2021).

3.	CORINE Land Cover — Copernicus Land Monitoring Service. https://land.copernicus.eu/pan-european/corine-land-cover.

4.	Landsat Missions. Landsat Normalized Difference Vegetation Index | U.S. Geological Survey. https://www.usgs.gov/landsat-missions/landsat-normalized-difference-vegetation-index.

5.	Landsat Missions. Landsat Modified Soil Adjusted Vegetation Index | U.S. Geological Survey. https://www.usgs.gov/landsat-missions/landsat-modified-soil-adjusted-vegetation-index.

6.	Sexton, J. O. *et al.* Earth Science Data Records of Global Forest Cover and Change User Guide 2 Global Land Cover Facility. (2016).

7.	European Settlement Map — Copernicus Land Monitoring Service. https://land.copernicus.eu/pan-european/GHSL/european-settlement-map.

8.	Imperviousness — Copernicus Land Monitoring Service. https://land.copernicus.eu/pan-european/high-resolution-layers/imperviousness.

9.	WorldPop: Population Counts. https://hub.worldpop.org/project/categories?id=3.

10.	*EU-DEM Statistical Validation*. https://land.copernicus.eu/user-corner/technical-library/eu-dem-2013-report-on-the-results-of-the-statistical-validation (2014).

11.	Tilastokeskus Esri Finland. Tilastokeskuksen tilastot postinumeroalueittain. Koordinaattijärjestelmä TM35FIN. https://www.arcgis.com/home/item.html?id=f1f6d6432a89424789c1fb13fba3951d (2022).
